# ASSESSMENT OF COAGULATION FACTORS IN PATIENTS OF SEVERE RHEUMATIC MITRAL STENOSIS IN SINUS RHYTHM WITH LEFT ATRIAL APPENDAGE INACTIVITY

**DOI:** 10.1101/2023.11.02.23298010

**Authors:** Saibal Mukhopadhyay, Narendra Kumar Chauhan, Sanjeev Kathuria, Bhawna Mahajan, Ghazi Muheeb, Vimal Mehta, Rupesh Santosh Agrawal, Sunil Kumar Mandal, Jamal Yusuf

## Abstract

**Background:** Patients of severe mitral stenosis (MS) in normal sinus rhythm (NSR) with left atrial appendage (LAA) inactivity and associated left atrial spontaneous echo contrast (LASEC) develop left atrial (LA) or LAA thrombus. But unlike atrial fibrillation (AF), oral anticoagulants (OAC) are not routinely prescribed in this subset of patients.

**Aim:** To assess local (LA) and systemic levels of procoagulants (PF1+2: prothrombin fragment 1+2; TAT-III: thrombin antithrombin III), PAI-1 (plasminogen activator inhibitor-1) and fibrinogen, in patients of severe MS in NSR with LAA inactivity and associated LASEC with healthy controls.

**Methods:** 35 patients of severe MS with valve suitable for balloon mitral valvuloplasty, along with 35 healthy volunteers were enrolled. All patients underwent transthoracic and transesophageal echocardiography to assess severity of MS, LAA activity, grade of LASEC, and exclude the presence of LA or LAA clot. Peripheral venous and LA blood samples were analysed for levels of procoagulants.

**Results:** Baseline characteristics like age and sex were comparable in both groups. Most of the patients in our study were either in NYHA II (n=13, 37.1%) or NYHA III functional class (n=21, 60%) and had grade 3+ (n=17;48.57%) or grade 4+ (n =15;42.86%) LASEC. Levels of PF1+2 {patient vs control, 9017(6228-10963.5) pg/mL vs 1790(842.3-2712) pg/mL, p<0.0001)}, TAT-III {patient vs control, 39(5.45-74.85) ng/ml vs 2.80(1.6-6.5) ng/mL, p<0.0001}, PAI-1 {patient vs control, 26.09±8.18 ng/ml vs 8.05 ± 3.53ng/ml. p<0.0001)}and fibrinogen (3.48± 0.89g/L vs 3.01± 0.53g/L, p=0.029) were significantly higher in LA of patients as compared to controls. Similarly, systemic levels of PF1+2, TAT-III, PAI-1 and fibrinogen were significantly higher in patients as compared to controls. However, systemic level of D-dimer was similar in both groups.

**Conclusion:** Both local and systemic levels of procoagulants were significantly raised in patients of severe MS in NSR with LAA inactivity and associated grade 3+ or 4+ LASEC, suggestive of a hypercoagulable state similar to that reported in patients of AF. Hence, we feel that OAC should be administered routinely in this subgroup of patients to prevent thrombus formation until there is improvement in LA and LAA function following valvuloplasty or mitral valve surgery.

**Clinical Perspective:** *What’s new?:*

- Patients of severe MS in NSR with LAA inactivity and associated LASEC are prone to develop LA or LAA thrombus.
- However, the ACC/AHA 2020 guidelines on valvular heart disease still do not recommend oral anticoagulants in this subset of patients.
- We carried out an adequately powered study to assess the level of procoagulants in patients of severe MS in NSR with associated LASEC (LASEC being a marker of stasis).

*What are the clinical implications?:*

- Both local and systemic levels of procoagulants were significantly raised in patients of severe MS in NSR with LAA inactivity and associated grade 3+ or 4+ LASEC as compared to controls, suggestive of a hypercoagulable state similar to that reported in patients of AF.
- We feel that OAC should be administered routinely in this subgroup of patients of rheumatic MS in NSR to prevent thrombus formation until there is improvement in LA and LAA function following valvuloplasty or MV surgery.

## Introduction

Thromboembolic events are a major cause of morbidity and mortality in patients of mitral stenosis (MS).^1,2^ While left atrial (LA)/left atrial appendage (LAA) thrombus formation has been reported in 33% patients of rheumatic MS in atrial fibrillation (AF),^3^ it has also been reported in 6.6-13.5% of patients in normal sinus rhythm (NSR).^4,5,6,7^

The LAA is the commonest site of thrombus formation. The stasis of blood flow in LAA, which occurs in the absence of forcible atrial contraction during AF, initiates the clot formation process and leads to the occurrence of arterial thromboembolic events.^7,8^ Serkan et al^6^ in an interesting study, has shown that in patients of severe MS, there is an impairment of LA/LAA contractility to a similar extent, irrespective of whether a patient is in sinus rhythm (SR) or AF. Impairment of LA/LAA contractility leads to stasis of blood with formation of smoke in LA/LAA (LASEC), which in turn has been found to be a positive predictor of LA/LAA thrombus in patients of severe rheumatic MS in NSR.^7,9,10,11^ A number of small studies have also reported that patients with severe rheumatic MS in NSR with associated LASEC have raised atrial and systemic levels of procoagulants.^9,12,13^

However, the ACC/AHA 2020 guidelines on valvular heart disease^14^ still do not recommend oral anticoagulants in patients of severe rheumatic MS in NSR with associated LASEC. As stasis of blood (LASEC being a marker of stasis) and hypercoagulable state are prerequisites for thrombus formation, we decided to carry out an adequately powered study to assess the level of procoagulants in patients of severe MS in NSR with associated LASEC.

## Material and Methods

This was a prospective observational study conducted from August 2021 to January 2023 at Govind Ballabh Pant Institute of Postgraduate Medical Education and Research (GIPMER), New Delhi. The study aimed to assess and compare the level of procoagulants between patients of severe rheumatic MS in NSR and healthy controls. The ethics committee at our institution approved this study, and all patients provided written informed consent.

### Primary and secondary objectives

The primary objective was to assess the level of procoagulant prothrombin fragment 1+2 (PF1+2) in the left atrium and peripheral blood of patients of rheumatic MS in NSR compared to healthy control. The secondary objectives were to assess the level of thrombin antithrombin III complex (TAT III) (byproduct of thrombin generation), plasminogen activator inhibitor-1 (PAI-1) (marker of fibrinolytic system activation), fibrinogen and D dimer compared to healthy controls.

### Sample Size

Based on a previous study by Atak et al^12^ the mean standard deviation (SD) of PF1+2 in left atrial blood of rheumatic MS patients in NSR and associated LASEC was 4050 (±2100) pg/mL and in systemic blood of healthy controls, it was 2020 (±1400) pg/mL. Considering 1500 pg/mL as a clinically important difference between the disease group and the healthy control group and assuming a standard deviation (SD) of 2100 pg/mL in both groups, a sample of 32 subjects in each arm was needed with a power of 80% and a 5% level of significance. Thus, for the present study, we recruited 35 patients and 35 healthy controls to reduce the margin of error.

### Patients

The participants in the study group were men or women with an age more than 18 years with severe rheumatic MS {mitral valve area (MVA) ≤ 1.5 cm^2^ by planimetry}, valve suitable for balloon mitral valvuloplasty (BMV) and associated LASEC with LAA inactivity (LAA emptying velocity; LAAEV < 25 cm/s).^15,16^

Patients were excluded from the study if they had LA/LAA thrombus on echocardiography, AF, coexisting left ventricular dysfunction (ejection fraction <50%), congenital heart disease, or any contraindication for transesophageal echocardiography (TEE). Patients were also excluded if they had diabetes mellitus, hypertension, renal or hepatic dysfunction, overt malignancy, pregnancy, chronic inflammatory disease such as systemic lupus erythematosus, rheumatic arthritis, human immunodeficiency virus infection, a history of systemic or pulmonary embolism, or an associated COVID-19 infection.

### Controls

35 healthy volunteers were taken as controls for the study. The controls had no echocardiographic evidence of structural heart disease, were in sinus rhythm, and had all other exclusion criteria identical to those of the study patients.

### Echocardiography

All patients underwent detailed transthoracic echocardiography (TTE) in order to evaluate valvular involvement in the study group and exclude cardiac disease in control subjects using a Philips EPIQ 7C fitted with a commercially available 5 MHz transducer. Two-dimensional and Doppler echocardiographic studies were performed in the left lateral decubitus position in conventional views (parasternal long, short-axis, and apical two- and four-chamber views) according to American Society of Echocardiography guidelines.^17^ A one lead electrocardiogram was recorded continuously during the TEE (informed written consent was taken from all patients before performing multiplane TEE) using a Philips EPIQ 7C fitted with a commercially available 5 MHz X7-2t transducer. The LA and LAA imaging were begun in the horizontal plane at 0°, then the transducer was rotated progressively to 60° and 90°, in the same plane, and images were also evaluated after a slight and more pronounced anticlockwise rotation of the probe. The rotation of the transducer to 110° and 130° was coupled with a more pronounced anticlockwise rotation of the probe. All images were recorded by optimizing the gain settings to minimize gray-noise artefacts. We examined the LAA on the short and long axis. It has been shown that TEE is highly sensitive for the detection of left atrial clots, especially in the left atrial appendage.^18^

A LA or LAA thrombus was diagnosed by the presence of a clearly defined echogenic intracavitary mass that was different from the underlying endocardium. Spontaneous Echo Contrast (SEC) was diagnosed by the presence of dynamic smoke-like echoes in the LA cavity and LAA with a characteristic swirling motion. Patients with LA/LAA thrombus were excluded from the study. The severity of SEC was graded from 0 to 4+, as proposed by Fatkin et al.^19^ Grade 0: None (absence of echogenicity); Grade 1+: Mild (minimal echogenicity located in the LA appendage or sparely distributed in the main cavity of the left atrium; may be detectable only transiently during the cardiac cycle; imperceptible at operating gain settings for two-dimensional echocardiographic analysis); Grade 2+: Mild to moderate (denser swirling pattern than grade 1+ but with similar distribution; detectable without increased gain settings); Grade 3+: Moderate (dense swirling pattern in the LAA, generally associated with somewhat lesser intensity in the main cavity; may fluctuate in intensity but be detectable constantly throughout the cardiac cycle); Grade 4+: Severe (intense echo density and very slow swirling patterns in the LAA, usually with similar density in the main cavity).

### LAA Velocity

The LAA flow profiles were obtained by placing the sample volume of the pulsed Doppler 1 cm below the LAA orifice, where there were no wall artefacts and a net flow could be recorded using pulse wave Doppler. The positive flow observed after the P-wave of the surface ECG was taken as LAAEV, representing the LAA contractile function. Inactive LAA was defined as late peak LAAEV <25 cm/s.^15,16^ As in our previous study,^20^ LAAEV was recorded in three consecutive cycles in two views (mid-esophageal short axis view and two chamber view). Only those patients with mean LAAEV values of <25 cm/s in both the views were enrolled for the study. All echocardiograms were independently evaluated by two observers, and any difference of opinion was settled by mutual consensus.

### Valvuloplasty Procedure

In the catheterization laboratory, introducer sheaths were inserted into the right femoral artery and vein. A 6F polymer pigtail catheter was inserted into the left ventricle for pressure measurement. A trans-septal puncture was performed with the help of a Brockenborough needle in a 6F Mullin sheath, and the mitral valve (MV) was dilated with an Inoue balloon.^21^ No thromboembolic events occurred in any patient during or after the valvuloplasty procedure.

### Blood Sampling

Prevalvuloplasty peripheral blood samples were drawn from a femoral vein sheath and immediately transferred to an EDTA tube containing 3.8% sodium citrate. Immediately after trans-septal puncture, left atrial prevalvuloplasty samples were withdrawn from all 35 patients through the transseptal catheter (Mullin sheath) before administration of heparin. In controls, peripheral venous blood samples were collected through a needle puncture after discarding the initial 3 mL of blood. Blood samples for procoagulants were immediately (within 2 hours) centrifuged at 3000 x g for 15 minutes, and the collected plasma was stored at -70°C in several aliquots in our biochemistry laboratory for assessment in a blinded manner.

### Hematological Investigations

Baseline laboratory tests [haemogram, erythrocyte segmentation rate, C reactive protein, renal function test, liver function test, PT (prothrombin time), aPTT (activated partial thromboplastin time), fibrinogen, and platelet count] were done in all subjects of our study (patients and controls).

### Assay Procedure

Peripheral venous blood sample of controls for measuring levels of plasma coagulation parameters, including PF1+2, TAT-III and PAI-1, was drawn in the morning between 8 and 10 a.m. Samples were taken using 21-G vacuum tube phlebotomy needles into 3.8% 1:9 trisodium citrate-containing tubes, without venous stasis. Both left atrial blood and femoral vein samples of patients were taken during BMV before administration of heparin and immediately transferred to 3.8% 1:9 trisodium citrate containing tubes. The plasma was separated by centrifugation of blood (3000 x g for 15 min), and stored in several aliquots at - 70°C until used for the assay.

During thrombin generation, the amino terminus of the prothrombin molecule is released as the inactive PF1+2. Once evolved, thrombin converts fibrinogen into fibrin, which crosslinks to form thrombus, thereby releasing fibrinopeptide A (FPA) and B (FPB), or it can be inhibited by the endogenous heparin sulfate-antithrombin III mechanism to form the TAT-III complex which is a stable enzyme-inhibitor. Thus, PF1+2 and TAT-III are the markers of thrombin generation.^22^

The fibrinolytic system consists of a series of activators and inhibitors that regulate the conversion of plasminogen to plasmin. The major activators of plasminogen are tissue type plasminogen activator and urokinase, both of which are inactivated by the circulating inhibitor, PAI. Freely circulating plasmin is inactivated by α2-plasmin inhibitor, forming a plasmin-α2-plasmin inhibitor complex. On the other hand, fibrin formed from fibrinogen is hydrolyzed by plasmin, releasing D-dimer. Thus, both plasmin-α2-plasmin inhibitor complex and D-dimer levels reflect fibrinolytic status, whereas PAI is a marker of impaired fibrinolysis.^22^

Plasma PF1+2 concentration, used as a marker of in vivo thrombin generation, was measured using a quantitative sandwich enzyme-linked immunosorbent assay method (ELISA KIT, Elabscience USA, normal human plasma values: 31.2–2000 pg/mL; intra- and inter-coefficients of variation < 10%). Plasma TAT-III concentration, also a marker of in vivo thrombin generation, was measured using a quantitative sandwich enzyme-linked immunosorbent assay method (ELISA KIT, Elabscience USA; normal human plasma values: 0.5–10 ng/ml; intra- and inter-coefficients of variation < 10%). PAI-1, used as a marker of endogenous fibrinolytic activity, was measured using a quantitative sandwich enzyme-linked immunosorbent assay (Elabscience, USA; normal plasma value is 5–40 ng/ml; intra- and inter-assay coefficients of variation are < 10%). Plasma fibrinogen was assayed by the direct chromogenic method. D-dimer was measured with a particle-enhanced immune-turbidimetric method. The analysis of plasma samples was done in the Biochemistry Department of our hospital in a blinded manner.

### Statistical Analysis

The categorical variables have been presented as numbers and percentages (%). On the other hand, the quantitative data have been presented as mean ± SD and as median with the 25^th^ and 75^th^ percentiles (interquartile range). The data’s normality was checked using the Kolmogorov-Smirnov test. In the cases in which the data was not normal, we used non-parametric tests. The comparison of the variables that were quantitative and not normally distributed in nature was analysed using the Mann-Whitney Test (for two groups), and the variables that were quantitative and normally distributed were analysed using the independent T test (for two groups). A paired T test/Wilcoxon signed rank test was used for comparison between peripheral blood and left atrial blood. The comparison of the variables, which were qualitative in nature, was analysed using the Chi-Square test. If any cell of a contingency table had an expected value of less than 5, then Fisher’s exact test was used.

The data entry was done in the Microsoft Excel spreadsheet, and the final analysis was done with the use of Statistical Package for Social Sciences (SPSS) software, IBM manufacturer, Chicago, USA, version 25.0. A p value of less than 0.05 was considered statistically significant.

## Results

The study was conducted from August 2021 to January 2023 at Govind Ballabh Pant Institute of Postgraduate Medical Education and Research (GIPMER), New Delhi, and enrolled 35 patients with severe rheumatic MS in NSR with associated LASEC and an equal number of healthy controls. No statistically significant differences were observed in terms of age, sex, or body mass index between the two study groups. The average age for both groups was 32 years, and approximately two-thirds of the subjects were female (Table 1). Hematological parameters and coagulation profiles were found to be comparable between the patient and control groups (Table 2).

**Table 1:** Baseline characteristics of the study population.

| Variables | MS in SR<br>(35) | Controls<br>(35) | p value |
| --- | --- | --- | --- |
| Age (years)<br>(Mean ± SD) | 32.24 ± 7.44 | 32.93 ± 2.64 | 0.791 |
| Sex (female)<br>n (%) | 24 (68.57%) | 23 (65.7%) | 0.799 |
| BMI (kg/m<sup>2</sup>) (Mean ± SD) | 22.01 ± 2.88 | 21.63 ± 2.25 | 0.739 |
| NYHA Functional Class |  |  |  |
| NYHA II | 13 (37.14%) |  |  |
| NYHA III | 21 (60%) |  |  |
| NYHA IV | 1 (2.87%) |  |  |

BMI= Body mass index, SR = Sinus rhythm, NYHA = New York Heart Association functional classification of heart failure.

**Table 2:** Comparison of hematological parameters between patients with mitral stenosis in sinus rhythm and controls.

| Variables | MS in SR<br>(35) | Controls<br>(35) | p value |
| --- | --- | --- | --- |
| Hb (g/dL) | 12.91 ± 1.62 | 13.03 ± 2.05 | 0.662 |
| Hematocrit (%) | 40.39 ± 4.37 | 40.95 ± 5.85 | 0.925 |
| Platelets count (Thousand/mm<sup>3</sup>) | 233.26 ± 73.58 | 256.63 ± 66.2 | 0.091 |
| SGOT (IU/L) | 24.8 ± 8.05 | 20.7 ± 6.17 | 0.020 |
| SGPT (IU/L) | 20.14 ± 9.99 | 19.07 ± 7.3 | 0.611 |
| Total bilirubin (mg/dL) | 0.67 ± 0.36 | 0.65 ± 0.24 | 0.218 |
| Total protein (g/dL) | 7.68 ± 0.55 | 7.57 ± 0.37 | 0.112 |
| CRP (mg/dL) | 1.1 (0.50-2.85) | 1.32 (0.904.10] | 0.211 |
| ESR (mm/hr) | 19.06 ± 10.48 | 24.84 ± 16.47 | 0.143 |
| Bleeding time (seconds)<br>(Mean ± SD) | 72 ± 17.54 | 72.87 ± 16.59 | 0.435 |
| Clotting time (seconds)<br>(Mean ± SD) | 591.57 ± 59.13 | 591.53 ± 86.42 | 0.987 |
| INR (Mean ± SD) | 0.92 ± 0.12 | 0.95 ± 0.1 | 0.790 |
| aPTT (seconds) (Mean ± SD) | 25.45 ± 5.56 | 25.85 ± 6.59 | 0.784 |

Hb = hemoglobin, SGOT = serum glutamic-oxaloacetate transaminase, SGPT = serum glutamate pyruvate transaminase, CRP = C reactive protein, ESR = erythrocyte sedimentation score, INR = international normalized ratio, aPTT = activated partial thromboplastin clotting time.

This study included patients diagnosed with severe mitral stenosis (MVA ≤1.5 cm^2^), and the mean MVA was 0.85 cm^2^ by planimetry. TEE revealed that the majority of patients exhibited grades 3+ (n = 17; 48.57%) or 4+ LASEC (n = 15; 42.86%)}. The mean LAAEV was 16.38±4.04 cm/s as all our patients had inactive LAA as an inclusion criterion. The mean gradient across the MV was 15.2 ± 5.1 mmHg, suggestive of severe MS (Table 3).

**Table 3:**
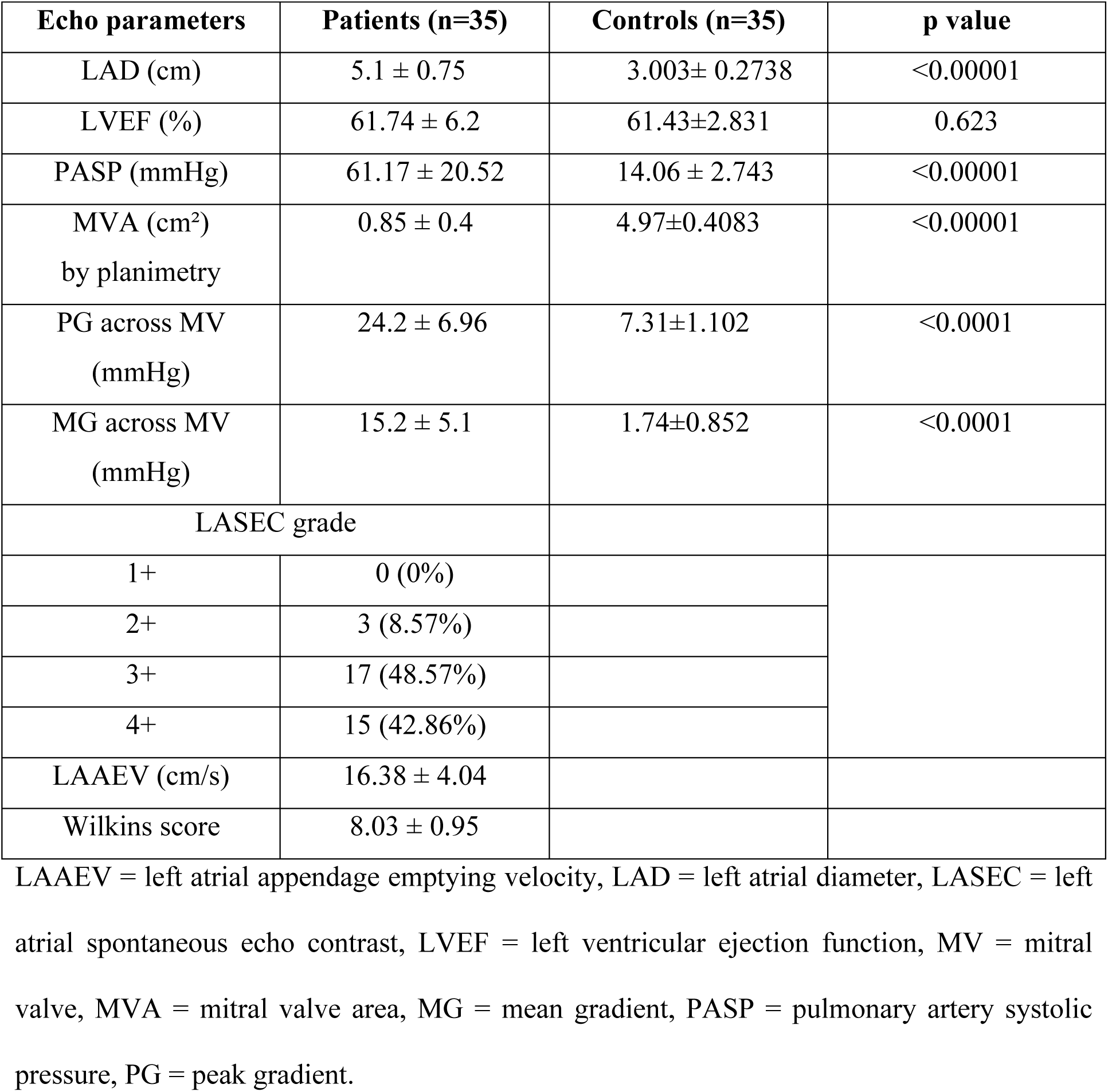
Echocardiographic parameters in patients of mitral stenosis in sinus rhythm and controls.

Significant elevations of procoagulants, specifically PF1+2 and TAT-III, were observed in patients as compared to controls. In the patient group, the systemic level of PF1+2 was significantly higher than that of healthy controls [(2931 (1626-7770) pg/mL vs 1790 (842.3-2712) pg/mL; p<0.0001)], indicating the presence of a systemic hypercoagulable state (Table 4). Moreover, the LA level of PF1+2 was significantly higher than the systemic level in patients, primarily attributed to increased activation of the coagulation system in LA compared to peripheral blood. Similarly, the systemic level of TAT-III was significantly higher in patients compared to controls [14.09 (4.9-42.725) ng/mL vs 2.80 (1.60-6.50) ng/mL; p<0.0001], and the LA level of TAT-III showed a significant elevation relative to the systemic levels in patients [39 (5.45-74.85) ng/mL vs 2.80 (1.60-6.50) ng/mL; p<0.0001) (Table 4).

**Table 4:** Comparison of coagulation factors and fibrinolytic system in patients of mitral stenosis and controls.

| Variable of dispersion | Peripheral blood of patients (n=35) | Left atrial blood of patients (n=35) | Peripheral blood of Controls (n=35) | p value (LA blood of patients vs peripheral blood of controls) | p value (peripheral blood of patients vs controls) | p value (peripheral blood vs LA blood of patients) |
| --- | --- | --- | --- | --- | --- | --- |
| PF1+2 (pg/mL) |  |  |  |  |  |  |
| Median (25<sup>th</sup>-75<sup>th</sup> percentile) | 2931 (1626-7770) | 9017 (6032-10963.5) | 1790 (842.3-2712) | <0.0001 | <0.0001 | <0.00001 |
| TAT-III (ng/mL) |  |  |  |  |  |  |
| Median (25<sup>th</sup>-75<sup>th</sup> percentile) | 14.09 (4.9-42.725) | 39 (5.45-74.85) | 2.80 (1.60-6.50) | <0.0001 | <0.0001 | <0.00001 |
| PAI-1 (ng/mL) |  |  |  |  |  |  |
| Mean ± SD | 15.11±6.74 | 26.09±8.18 | 8.05±3.53 | <0.0001 | 0.001 | <0.001 |
| Plasma fibrinogen (g/L) |  |  |  |  |  |  |
| Mean | 3.91±1.29 | 3.48±0.89 | 3.01±0.53 | 0.029 | 0.001 | 0.53 |
| D dimer (ng/mL) |  |  |  |  |  |  |
| Median (25<sup>th</sup>-75<sup>th</sup> percentile) | 430 (230-690) | 290 (110-610) | 250 (160-460) | 0.061 | 0.737 | 0.485 |

PF1+2 (prothrombin fragment 1+2), PAI-1 (plasma activator inhibitor), TAT-III- Thrombin antithrombin III complex.

In addition to the observed increase in thrombin generation in the LA, the patient group also displayed impairment of the fibrinolytic system. The systemic level of PAI-1 was elevated in patients with MS compared to healthy controls, indicative of systemic impaired fibrinolytic activity. Furthermore, the LA level of PAI-1 was significantly higher compared to the systemic level, suggesting a significant hypercoagulable state in the LA compared to the periphery in the patient group [26.09±8.18 ng/mL vs 8.05±3.53 ng/mL; p<0.0001] (Table 4). Accordingly, the level of D-dimer (the end product of fibrin degeneration) was not significantly different in the LA of patients compared to controls, suggesting a significant impairment of the fibrinolytic system in this subset of patients (Table 4).

The mean plasma fibrinogen concentration in the peripheral blood of the patients was 3.91±1.29 g/L, while in the LA, it was 3.48±0.89 g/L. In comparison, the control group exhibited a mean fibrinogen level of 3.01±0.53 g/L. Statistical analysis revealed no significant disparity between systemic and LA fibrinogen levels in the patient group (p = 0.104). Nevertheless, these levels were notably elevated when compared to the healthy control group, suggesting the presence of a hypercoagulable state in patients of MS in SR (Table 4).

## Discussion

To the best of our knowledge, this is the first adequately powered study that has systematically evaluated the level of procoagulants in the left atrium of patients with severe rheumatic MS in sinus rhythm with LAA inactivity and associated LASEC compared to those of healthy controls.

As per Virchow’s triad,^23,24,25^ the three factors required for the formation of a thrombus in a vessel are:

i. endothelial injury,
ii. local blood stasis and
iii. hypercoagulable state.

All three of the above factors are responsible for the formation of thrombus in patients of MS in AF. In all patients of MS with AF, injury to the endothelium in the left atrium has been documented.^26^ Onset of AF leads to gross impairment of contractility of both LA and LAA, leading to stasis of blood, which in turn leads to crosslinking of RBCs with fibrin, forming smoke in LA/LAA.^27^ This LA/LAA smoke in turn leads to the activation of the coagulation system, creating a hypercoagulable milieu in the left atrium and thrombus formation.^9,28,29^ Accordingly, to prevent thrombus formation, the latest ACC/AHA guidelines on valvular heart diseases ^14^ recommend oral anticoagulants to all patients of MS with AF to prevent thrombus formation in LA and its devastating sequelae of systemic thromboembolism.

The reported incidence of LA/LAA clots in patients of MS in sinus rhythm has been reported to be 6.6-13.5% primarily affecting the young population in our country.^4,5,6,7^ As in patients of MS in AF, a similar degree of left atrial endocardial injury has been documented by electron microscopy in patients of MS in sinus rhythm.^26^ Secondly, LAA hypocontractility has been documented in 73% of patients of severe MS in sinus rhythm by us. We have also shown that LAA inactivity is the only independent predictor of LASEC, a marker of LA/LAA stasis in this subset of patients.^20^

Thirdly, a large number of small observational studies have also reported a hypercoagulable state in the left atrium of patients of MS in sinus rhythm.^9,12.28^ Peverill et al^9^ reported that LASEC is the only independent predictor of increased local activity of the coagulation system, irrespective of whether patients are in SR or AF. They also showed that the local level of procoagulant (PF1+2) was similar in patients of MS in AF (n=9) to those in sinus rhythm with LASEC (n=16).

In our study, we enrolled patients of severe MS with associated LASEC because this subset of patients has both local^9,12^ and systemic hypercoagulable state^9,12,13^ as described in patients with AF.^30,31,32^ Atak et al^12^ reported that the left atrial level of PF1+2 was significantly higher in patients of MS in SR, irrespective of LASEC. However, the peripheral level of PF1+2 was significantly increased only in LASEC positive patients compared to controls. Similarly, Ileri et al^13^ reported that peripheral level PF1+2 was significantly raised only in patients of MS in SR with LASEC. Peverill et al^9^ also reported that plasma levels of PF1+2 in both left atrial and peripheral blood were significantly higher in patients of MS in sinus rhythm with LASEC than in patients without LASEC.

The latest ACC/AHA guidelines on valvular heart disease^14^ still do not recommend oral anticoagulants in patients of severe MS in sinus rhythm with LASEC based on the study by Manjunath et al.^4^ In this cross-sectional study they enrolled 848 patients with rheumatic MS in SR to determine the incidence of LA clot and establish the factors, that play a determining role in this process. 6.6% patients in this study had LA/LAA clot. They found that LASEC is not an independent predictor of LA thrombus formation though all their patients with clot had associated LASEC. The age of patients in their study was higher suggestive of longer duration of stasis. But patients with enlarged LA also develop asymptomatic episodes of AF and older patients may have had greater number of episodes producing a greater hypercoagulable milieu. Manjunath et al have mentioned this fact as a limitation in their study and they had not assessed local and systemic levels of procoagulants. A comparison between this subgroup of patients with LA/LAA clot of Manjunath et al and our study is shown in table 5. The MVA (0.85±0.4 cm^2^ vs 0.78±0.18 cm^2^, p = 0.2572), LA transverse diameter (5.1±0.75 cm vs 5.1±0.6 cm, p = 1.0), mean LAAEV (16.38±4.04 cm/s vs 17.9±10.5 cm/s, p = 0.3206) and grade of LASEC was similar in our study cases compared to Manjunath’s patients with LA/LAA clot. However, age, mean gradient across the MV and Wilkins score was higher in patients of Manjunath et al compared to our study.^4^ As MVA was similar in both the groups, higher transmitral gradient was probably due to significant subvalvular disease as was also shown by higher Wilkins score. The main effect of transmitral gradient is increased afterload on LA/LAA leading to impairment of its contractility and promoting stasis. In our previous study,^20^ we have already shown that mean gradient across MV ≥11 mmHg is an independent predictor of LAA stasis and in both the patient groups it was higher.

**Table 5:** Comparison of parameters between Manjunath clot group with our study.

| Characteristics | Manjunath clot group (n=56) | Our study (n=35) | p value |
| --- | --- | --- | --- |
| 1. Age (years) | 44.4±8.8 | 32.24±7.44 | <0.0001 |
| 2. MVA (cm<sup>2</sup>) | 0.78±0.18 | 0.85±0.4 | 0.2572 |
| 3. MG across MV (mmHg) | 18.5±6.0 | 15.2±5.1 | 0.0083 |
| 4. LASEC | 3.3±1.1 | 3.34±0.626 | 0.8451 |
| 5. LAAEV (cm/s) | 17.9±10.5 | 16.38±4.04 | 0.3206 |
| 6. LA transverse diameter (cm) | 5.1±0.6 | 5.1±0.75 | 1.00 |
| 7. Wilkins score | 9.4±1.2 | 8.03±0.95 | 0.0001 |

LA: left atrium; LASEC: left atrial spontaneous echo contrast; LAAEV: left atrial appendage
emptying velocity; MG; mean gradient; MV: mitral valve; MVA: mitral valve area.

The 2021 ESC guidelines for the management of valvular heart disease^33^ have also recommended oral anticoagulants in subset of patients of severe MS with dense LASEC or enlarged LA (transverse diameter > 5 cm) and in our study we have shown significantly elevated level of procoagulants in patients with both these features.

As this subset of patients of MS in NSR with grade 3+ or 4+ LASEC have a significantly high level of procoagulants in the LA/LAA together with a systemic hypercoagulable state, we feel that they should be considered for oral anticoagulants to prevent thrombus formation. Unlike patients of MS with AF who require lifelong anticoagulants, we feel that oral anticoagulants should be given to all patients of MS in SR with grade 3+ or 4+ LASEC until the local hypercoagulable milieu is altered by relieving the obstruction across the MV. Studies have shown that relieving the obstruction across the MV by valvuloplasty decreases the left atrial afterload, leading to improvements in LA/LAA contractility,^34,35^ decrease in stasis^36,37^ and decrease in the level of procoagulants.^38,39^

Hence, based on our study results, we feel that OAC should be routinely given to all patients of severe MS in NSR with grade 3+ or 4+ LASEC until there is improvement in LA/LAA function following valvuloplasty or MV surgery.

## Conclusion

Both local and systemic levels of procoagulants were significantly raised in patients of severe MS in NSR with LAA inactivity and associated grade 3+ or 4+ LASEC as compared to controls, suggestive of a hypercoagulable state similar to that reported in patients of AF. Hence, we feel that OAC should be administered routinely in this subgroup of patients to prevent thrombus formation until there is improvement in LA and LAA function following valvuloplasty or MV surgery.

## Data Availability

The authors confirm that the data supporting the findings of this study are available within the article.

## Acknowledgements

We are thankful to Dr. Rajeev Kumar Malhotra, scientist (statistician), Delhi Cancer Registry, DR. B.R.A. IRCH, AIIMS, Delhi for statistical analysis.

